# Differential Adaptive Immune Responses Following SARS-CoV-2 Infection in Children Compared to Adults

**DOI:** 10.1101/2024.04.05.24305283

**Authors:** Sabryna Nantel, Corey Arnold, Maala Bhatt, Yannick Galipeau, Benoîte Bourdin, Jennifer Bowes, Roger L. Zemek, Marc-André Langlois, Caroline Quach, Hélène Decaluwe, Anne Pham-Huy

**Affiliations:** Sainte-Justine University Hospital and Research Center, Montréal, Québec, Canada; Department of Microbiology, Infectious Diseases and Immunology, Faculty of Medicine, University of Montréal, Montréal, Québec, Canada; Department of Biochemistry, Microbiology and Immunology, Faculty of Medicine, University of Ottawa, Ottawa, Ontario, Canada; Children’s Hospital of Eastern Ontario Research Institute, University of Ottawa, Ottawa, Ontario, Canada; Division of Emergency Medicine, Department of Pediatrics, University of Ottawa, Ottawa, Ontario, Canada; Pediatric Immunology and Rheumatology Division, Department of Pediatrics, University of Montréal, Montréal, Québec, Canada; Division of Infectious Diseases, Immunology and Allergy, Department of Pediatrics, Children’s Hospital of Eastern Ontario, University of Ottawa, Ottawa, Ontario, Canada

**Author notes:** CORRESPONDING AUTHORS. Hélène Decaluwe, CHU Sainte-Justine Research Center, 3175, Chemin de la Côte-Sainte-Catherine, Montréal, QC, Canada (H3T 1C5), Anne Pham-Huy, Children’s Hospital of Eastern Ontario, 401, Smyth Road, Ottawa, ON, Canada (K1H 8L1). Shared senior authorships (Hélène Decaluwe, Anne Pham-Huy).

**Keywords:** SARS-CoV-2, children, T-cells, antibodies, human coronaviruses

## Abstract

**Background:** SARS-CoV-2 infection elicits distinct clinical features in children and adults. Profiling the adaptive immune response following infection in children is essential to better understand and characterize these differences.

**Methods:** Humoral and cell-mediated immune responses from unvaccinated pediatric and adult participants were analyzed following asymptomatic or mild non-Omicron SARS-CoV-2 infection. Levels of IgG and IgA targeting spike (S), receptor-binding domain (RBD), and nucleocapsid (N) proteins of SARS-CoV-2 were measured, while neutralizing antibody (nAb) titers were assessed against three viral strains (Wuhan, Omicron BA.1 and BA.4/BA.5). Specific T-cell memory responses were investigated by quantifying interferon-gamma (IFN-γ) secreting cells after stimulation with ancestral and variant strains of SARS-CoV-2, and seasonal human β- coronaviruses (HCoV)-OC43 and -HKU1.

**Results:** The study comprised 28 children (3 to 17 [median=10] years old) and 28 adults (19 to 62 [median=42]). At a mean time of seven months (± 2.8 months) after SARS-CoV-2 infection, children and adults mounted comparable antibody levels against S and RBD, as well as similar neutralization capacity. However, children displayed a weaker cellular memory response to SARS-CoV-2 than adults, with a median of 88 [28-184] spot forming units per million of PBMCs in children compared to 208 [141-340] in adults (***, P < .001). In children, the level of IFN-γ secreting cells in response to SARS-CoV-2 corresponds to that of seasonal coronaviruses.

**Conclusion:** Long-term memory T-cell responses to SARS-CoV-2 are enhanced in adults compared to children who demonstrate equivalent responses to SARS-CoV-2 and other HCoV.

**HIGHLIGHTS:**

- Children infected with SARS-CoV-2 show comparable binding and neutralizing antibody levels as adults seven months after infection.
- There are notable differences in the intensity of the T-cell response following SARS-CoV- 2 infection between children and adults.
- Children have more pronounced T-cell immunodominance towards the spike versus non- spike proteins compared to adults at seven months post-infection
- In contrast, T-cell responses to SARS-CoV-2 are globally reduced in children compared to adults but are alike to other seasonal β-coronaviruses.

## INTRODUCTION

Since the beginning of the COVID-19 pandemic, there has been age-related differences in clinical presentations and outcomes of SARS-CoV-2 infection. Severe disease disproportionately impacts older adults, in contrast to children, who generally have mild or asymptomatic disease.^1–3^ Several factors could account for these differences, such as fewer co-morbidities in children, frequency of exposure to SARS-CoV-2 and seasonal coronaviruses, differential expression of angiotensin-converting enzyme 2 (ACE2), which is required for viral entry, and host immune responses.^4–8^ Compared to adults, immunological studies following SARS-CoV-2 infection in children remain limited.^9,10^ Many studies have focused on humoral immunity following SARS-CoV-2 infection.^10,11^ Distinct antibody immune response dynamics are suggested in children compared to adults, but some results are conflicting between studies and many questions remain.^10,12,13^ Cell-mediated immunity is more complex to study and less has been published on the role of T cells in the acute and convalescent phases of infection in children, despite its importance in antiviral immunity and vaccine development.^14–17^

The goal of this study was to describe and compare the long-term adaptive immunity following SARS-CoV-2 infection in children and adults. To better understand the differences in memory immune responses following infection, humoral immunity was assessed through direct ELISA to measure binding antibody levels and surrogate neutralization ELISA to establish neutralizing antibody (nAb) titers. Cell-mediated immune responses were measured through ELISpot assay for the detection of functional antigen-specific T cells.

## METHODS

### Study Design and Data Collection

This study was a multicenter, longitudinal study, combining two existing prospective cohorts. Pediatric participants were recruited from the *Persistence of Antibody Titers to COVID-19 in Households (PATCH Study)*, a case-ascertained antibody-surveillance initiative based in Ottawa, Ontario.^18^ Children or adolescents with confirmed SARS-CoV-2 infection defined by either positive testing through PCR or positive anti-nucleocapsid (anti-N) SARS-CoV-2 IgG from the PATCH cohort were eligible to participate (n=38). Adult participants were selected from the *RE- Infection in COVID-19 Estimation of Risk Cohort (RECOVER Study)*, which consisted of health care workers who were recruited following PCR-confirmed SARS-CoV-2 infection (n=569).^19,20^ Adult samples were matched to the pediatric samples based on sex, ethnicity, and time since infection. All selected samples were from unvaccinated participants. Our study included one-time point per participant, which was around seven months post-infection. Questionnaires documented demographics, medical history, COVID-19 symptoms, and potential recent or recurrent SARS-CoV-2 infection.^18–20^

### Sample Collection and Processing

Blood samples from the pediatric cohort were collected at the Children’s Hospital of Eastern Ontario (CHEO) and cryopreserved at the Coronavirus Variants Rapid Response Network (CoVaRR-Net) Biobank within four hours (and up to 24 hours) of procurement, while those from the adult cohort were processed and cryopreserved at the Mother Child Biobank at the Sainte-Justine University Hospital and Research Center (CR-CHUSJ). Serum and peripheral blood mononuclear cells (PBMCs) were isolated for the assessment of antibody levels and T-cell studies, respectively. This was done according to standard operating procedures (SOPs) using SepMate^TM^ tubes (Stemcell Technologies, Canada). PBMCs were resuspended in complete Roswell Park Memorial Institute (RPMI) media (Gibco) with 10% dimethyl sulfoxide (DMSO) and cryopreserved in liquid nitrogen, while serum aliquots were cryopreserved at -80°C. Frozen samples from both cohorts were sent and analyzed using the same ELISpot assays (Dr. Decaluwe’s laboratory) and antibody and neutralization assays (University of Ottawa’s Serology and Diagnostics High-Throughput Facility, Dr. Langlois’ laboratory).

### Date of Infection

The date of infection was determined by the earliest of either 1) the onset of symptoms, or 2) the confirmed testing date. For asymptomatic individuals who were never tested but had confirmed infection through the presence of anti-N IgG, the infection date was estimated based on symptom onset or testing date of the first-infected member of the household.

### Study Period and SARS-CoV-2 Variant Circulation

Blood samples were collected from May to December 2021. The initial SARS-CoV-2 infections occurred between December 2020 and April 2021, before the emergence of the Omicron (BA.1) variant. Specific testing to identify SARS-CoV-2 variants was not conducted. Instead, the likely strain of SARS-CoV-2 was determined based on prevalent local epidemiology. Infections from March 2020 to March 2021 were most likely caused by the ancestral (Wuhan-like) strain, while those from March 2021 to April 2021 were predominantly attributed to the Alpha (B.1.1.7) variant.^21^

### Ethic Approvals

The study was approved by the Research Ethics Boards at the CHEO (**CHEOREB#21/08X**) and the CR-CHUSJ (**MP-21-2021-3035** and **MP-21-2021-3046)**. Consent, and assent when indicated, were obtained from all participants and/or their guardian.

### Humoral Immunity

*Chemiluminescent direct enzyme-linked immunosorbent assay (ELISA) for antibody detection* Serum samples were analyzed using automated chemiluminescent ELISAs for SARS-CoV-2- specific IgA and IgG targeting the whole spike (S) protein, the receptor binding domain of S (RBD), and the nucleocapsid (N) protein. This analysis was conducted with the Hamilton MicroLab STAR robotic liquid handlers at the University of Ottawa’s Serology and Diagnostics High-Throughput Facility (Faculty of Medicine), as previously described.^22^ Briefly, diluted serum samples were incubated in 384-well assay plates (Thermo Fisher Scientific, Waltham, MA, USA) coated with 50 ng/well of S, RBD or N (S and N from NRC Metrology Division, RBD from the laboratory of Dr. Yves Durocher). Secondary HRP-conjugated antibodies against IgG and IgA were added to the wells, and detected following incubation using SuperSignal Pico chemiluminescent substrate (Thermo Fisher Scientific, Waltham, MA, USA). Plate reader raw luminescence values were measured, blank-subtracted and scaled to an on-plate standard curve. Scaled values were then converted to international binding antibody units (BAU/mL) based on the WHO international standard (NIBSC code 20/136) and to lab-specific units (µg/mL) using established four-parameters logarithmic conversion models.^22^ For each antigen (S, RBD, and N), a cut-off was established based on false discovery rates in a pre-pandemic cohort (3%, 2%, and 5% respectively).^22,23^

### Chemiluminescent surrogate neutralization ELISA for nAb detection

Surrogate neutralization ELISA (snELISA) assesses the capacity of serum samples to inhibit the interaction between the S protein of SARS-CoV-2 and the ACE2 cell receptor.^22^ 384-wells assay plates were coated with 100 ng/well of trimeric S protein for ancestral SARS-CoV-2 (Wuhan, NRC Metrology Division) or the Omicron variant strains BA.1 and BA.4/5 (from the laboratory of Dr. Yves Durocher). Samples were serially diluted to generate five data points and incubated on the assay plates. Following incubation, 6.5 ng of biotinylated ACE2 (NRC Metrology Division, Nova Scotia, Canada) was incubated in each well, and subsequently detected using a streptavidin-peroxidase polymer (Thermo Fisher Scientific, Waltham, MA, USA) and SuperSignal Pico chemiluminescent substrate (Thermo Fisher Scientific, Waltham, MA, USA). Luminescence readings were background-subtracted, and each dilution point was expressed as a percentage of the average signal from wells with uninhibited antigen-ACE2 interaction (maximum signal). Three-parameter logarithmic regression was applied to model the % of inhibition as a function of the dilution factor, with fixed upper and lower points at 100% and 0%, respectively. The inhibitory dilution at 50% inhibition (ID50) for each sample was determined from its regression and presented as the reciprocal dilution factor (an ID50 of 100 indicates a sample diluted 1:100 has the capacity to inhibit 50% of ACE2-antigen binding).

### Cell-Mediated Immunity

#### Enzyme-linked immunospot (ELISpot) for the quantification of functional cellular immunity

Interferon-gamma (IFN-γ) secreting cells were detected through ELISpot assay. After rapid thawing and overnight resting, PBMCs were stimulated with one µg/mL of spike (S), nucleocapsid (N) or membrane (VME1) mega pools of SARS-CoV-2 peptides from the ancestral strain, as well as the spike from variants B.1.1.7 (Alpha), B.1.351 (Beta), P.1 (Gamma), B.1.617.2 (Delta), and B.1.1.529 (Omicron) and from two other β−coronaviruses (HCoV-OC43 and HCoV-HKU1) (JPT Peptide Technologies, JPT, Berlin, Germany) as described elsewhere.^24^ Culture media without peptide (AIM-V® Medium (1X), Thermo Fisher Scientific, Waltham, MA, USA) and CytoStim^TM^ (Miltenyi Biotec, MA, USA) were used as negative and positive controls respectively. Spots were quantified using CTL ImmunoSpot® SS UV Analyzer (Cellular Technology Ltd., OH, USA). The positive threshold response was defined as 25 spot-forming units (SFU) per million of PBMCs, as previously published by our team.^19,25^

#### Statistics

Mann-Whitney or Kruskal-Wallis unpaired nonparametric tests were performed using Prism 9, version 9.2.0 (2021 GraphPad Software, LLC) to assess statistical significance. Significance was set as *P <.05, **P <.01, ***P <.001 and ****P <.0001. Time since infection is presented as mean ± SD, while experimental data are displayed as median [interquartile range, 25^th^ – 75^th^ percentile].

## RESULTS

### Participant Characteristics

There were 28 children (median age = 10 years old [Min-Max, 3-17], 50 % female) and 28 adults (42 years old [19-62], 50 % female) included in this study for which samples were suitable for analysis (**Table 1**). One time point was analyzed per study participant, with a mean time since infection of 7.4 ± 2.8 months for children and 7.0 ± 2.5 months for adults. Most children reported mild COVID-19 symptoms, categorized as WHO scale 2-3, similar to the adult group, with the exception of four children who were asymptomatic and identified through household exposure history (**Table 1**). All children and 46.4% (n=13) of the adult cohort were presumed to be infected with the ancestral (Wuhan-like) strain as it was the predominant circulating strain at the time of infection. The remaining adults (n=15) were likely infected by the B.1.1.7 Alpha variant.

**Table 1.**
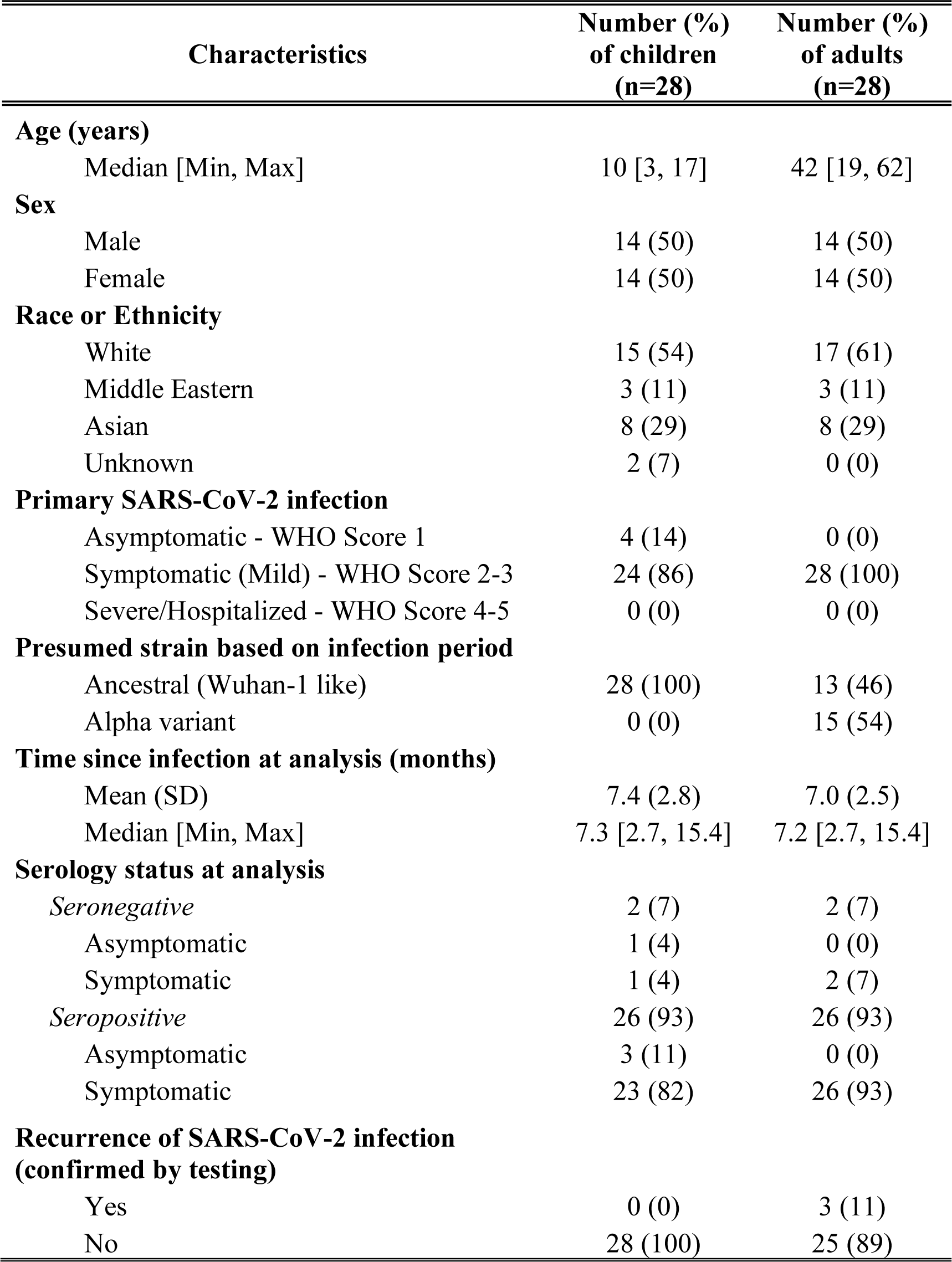
Characteristics of participants included in the analysis.

| Characteristics | Number (%)<br>of children<br>(n=28) | Number (%)<br>of adults<br>(n=28) |
| --- | --- | --- |
| **Age (years)** |  |  |
| Median [Min, Max] | 10 [3, 17] | 42 [19, 62] |
| **Sex** |  |  |
| Male | 14 (50) | 14 (50) |
| Female | 14 (50) | 14 (50) |
| **Race or Ethnicity** |  |  |
| White | 15 (54) | 17 (61) |
| Middle Eastern | 3 (11) | 3 (11) |
| Asian | 8 (29) | 8 (29) |
| Unknown | 2 (7) | 0 (0) |
| **Primary SARS-CoV-2 infection** |  |  |
| Asymptomatic - WHO Score 1 | 4 (14) | 0 (0) |
| Symptomatic (Mild) - WHO Score 2-3 | 24 (86) | 28 (100) |
| Severe/Hospitalized - WHO Score 4-5 | 0 (0) | 0 (0) |
| **Presumed strain based on infection period** |  |  |
| Ancestral (Wuhan-1 like) | 28 (100) | 13 (46) |
| Alpha variant | 0 (0) | 15 (54) |
| **Time since infection at analysis (months)** |  |  |
| Mean (SD) | 7.4 (2.8) | 7.0 (2.5) |
| Median [Min, Max] | 7.3 [2.7, 15.4] | 7.2 [2.7, 15.4] |
| **Serology status at analysis** |  |  |
| *Seronegative* | 2 (7) | 2 (7) |
| Asymptomatic | 1 (4) | 0 (0) |
| Symptomatic | 1 (4) | 2 (7) |
| *Seropositive* | 26 (93) | 26 (93) |
| Asymptomatic | 3 (11) | 0 (0) |
| Symptomatic | 23 (82) | 26 (93) |
| **Recurrence of SARS-CoV-2 infection**<br>**(confirmed by testing)** |  |  |
| Yes | 0 (0) | 3 (11) |
| No | 28 (100) | 25 (89) |

### Children show comparable levels of anti-spike antibodies but reduced anti-nucleocapsid antibodies when compared to adults

Seropositivity to the virus long after infection was assessed using SARS-CoV-2-specific IgG ELISA in both children and adult serum (**Table 1**). Most subjects were seropositive (93% in each cohort) at the time of immune response analysis. Children and adults had comparable levels of anti-S and anti-RBD antibodies, but children had lower antibody levels against non-spike components of SARS-CoV-2 (**Fig. 1**). Notably, both IgG (**Fig. 1A**) and IgA (**Fig. 1B**) binding to the N protein were significantly lower in children than in adults with a median of 31.0 [20.2-53.8] BAU/mL in children compared to 56.1 [32.6-113.8] BAU/mL in adults (*P < .05) for IgG and 0.02 [0.00-0.31] µg/mL in children compared to 0.29 [0.11-0.92] µg/mL in adults (**P < .01) for IgA. Because we previously reported that symptomatology and serology status affected the strength of the immune response in infected adults, we questioned if it may influence results in the pediatric population as well.^19^ When asymptomatic (n=4) and seronegative (n=1) children were excluded from the analysis, the difference in IgA levels against the nucleocapsid remained, while it was less evident for anti-N IgG levels (**Fig. S1**).

**Figure 1.**
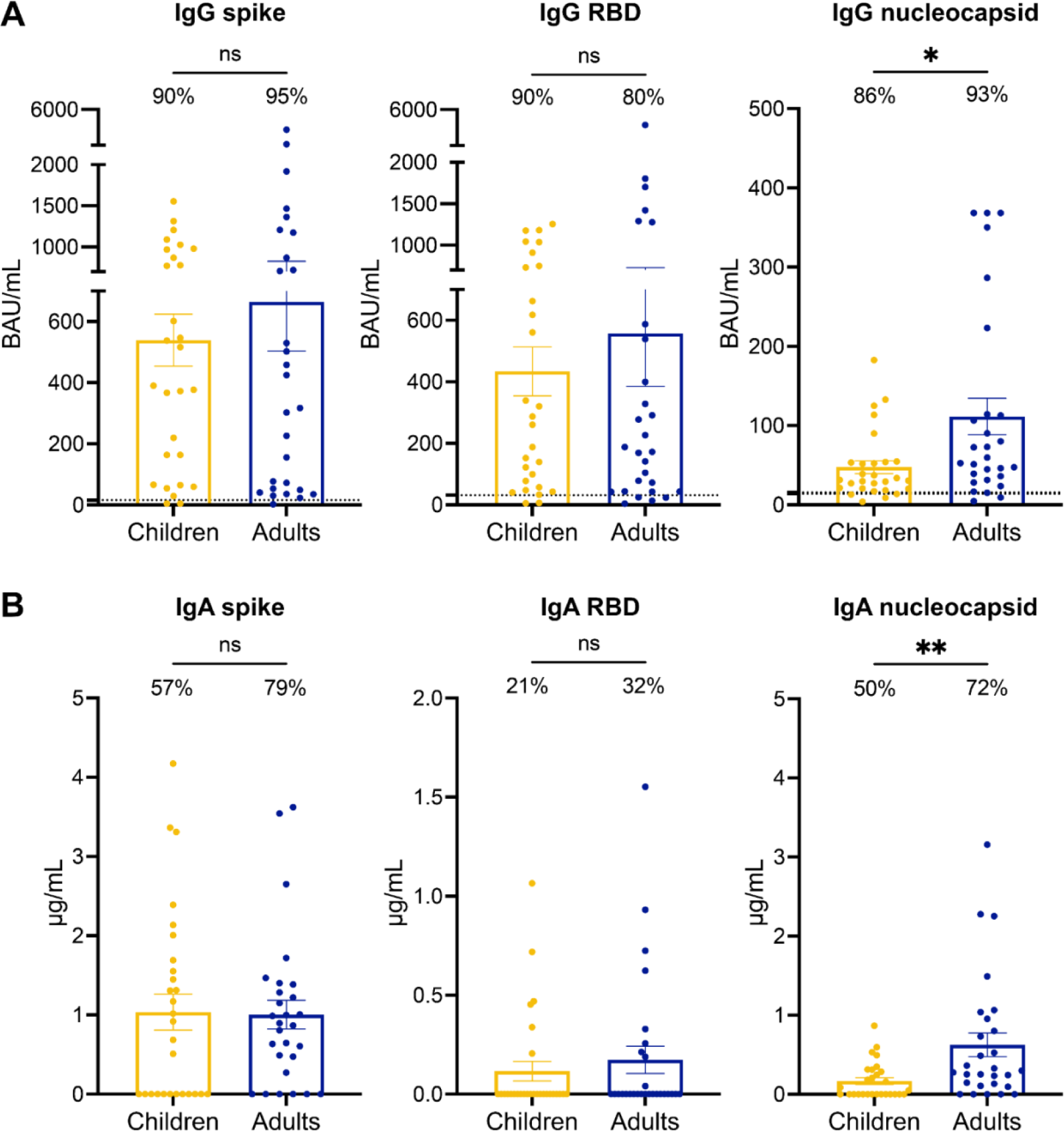
Children develop lower antibody levels against non-spike components of SARS-CoV-2 compared to adults. ELISA were conducted to measure the anti-spike **(left panels)**, anti-RBD **(middle panels)** and anti-nucleocapsid **(right panels)** IgG **(A)** and IgA **(B)** levels in the serum of children (n = 28, yellow) and adults (n = 28, blue) infected with SARS-CoV-2. **(A)** The percentage of participants with IgG responses above the positive cut-off value (15.53216 for spike, 31.34807 for RBD and 13.83556 for nucleocapsid) are indicated for each group. **(B)** The percentage of participants with detectable IgA are indicated for each group. Error bars indicate mean ± SEM. Statistical significance was established as : ns (not significant) P >.05, *P <.05, **P <.01.

### Children and adults exhibit comparable neutralizing capacity

Blockade of SARS-CoV-2 viral entry through the ACE2 receptor is largely mediated by immunoglobulins targeting the spike protein and its RBD component.^26–28^ Here, we assessed neutralizing capacity more than six months post-infection through surrogate neutralization ELISA against the ancestral SARS-CoV-2 as well as Omicron variants BA.1 and BA.4/BA.5. Children and adults had similar neutralizing capacity against all three strains assessed. As demonstrated in other studies, neutralizing capacity decreased against newer variants (Omicron BA.1 and BA.4/BA.5) compared to the ancestral strain, a finding equivalent in children and adults (**Fig. 2**).^29^ When analyzing only participants with symptomatic SARS-CoV-2 infection, we noted comparable neutralizing antibody levels for ancestral and Omicron BA.1, but slightly increased nAb titers against Omicron BA.4/BA.5 in children, with a median nAb titer of 2.58 [1.08-8.25] compared to 0.52 [0.07-3.72] in adults (*P < .05) (**Fig. S2**).

**Figure 2.**
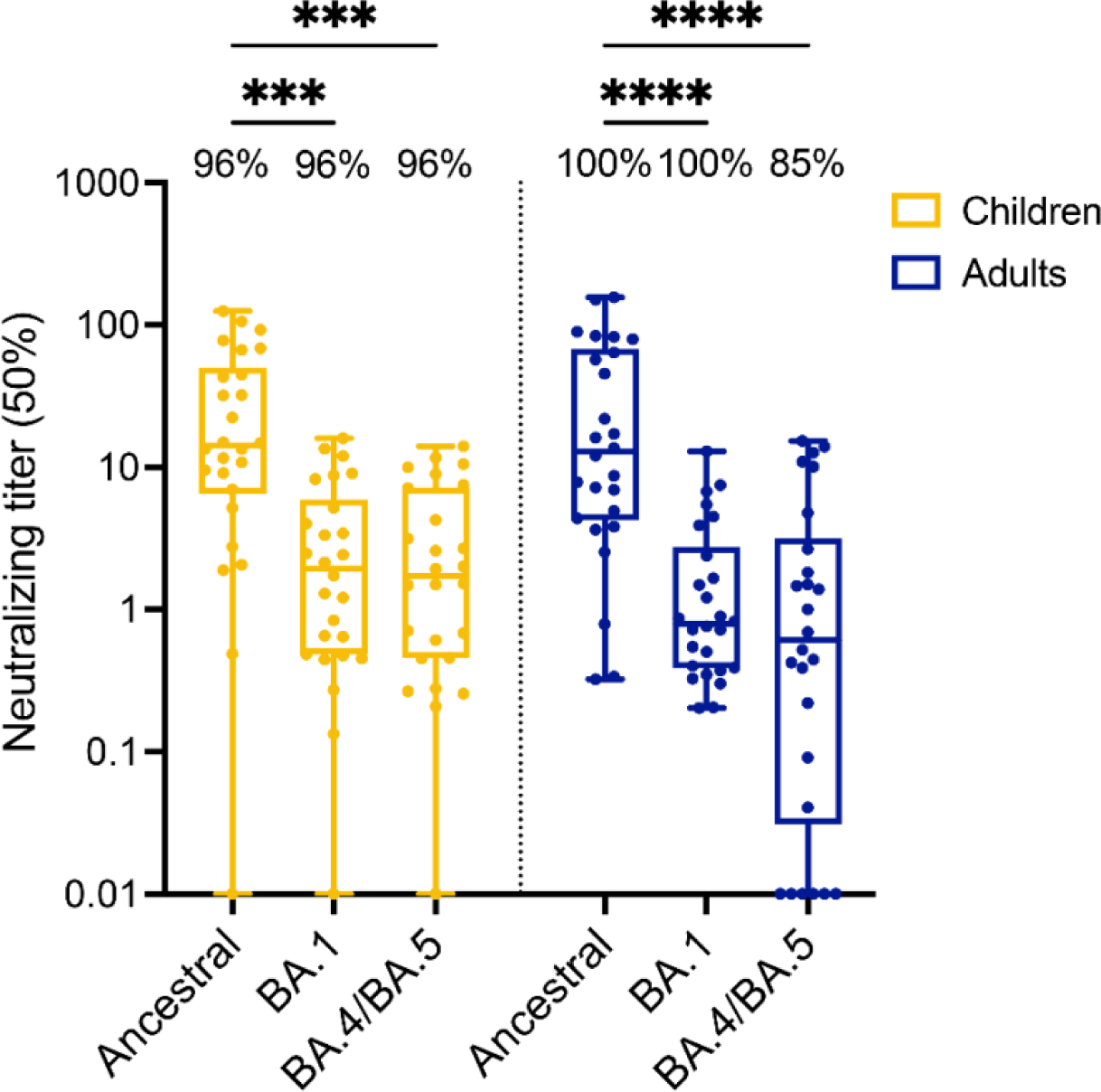
Children develop similar neutralizing capacity to adults after infection with SARS-CoV-2. Surrogate neutralization ELISA were conducted to establish the serum dilution level (ID50) required to inhibit 50% of the binding between the trimeric spike protein and the ACE2 receptor, thus neutralizing the virus attachment capability to cells. Neutralizing antibody titers were measured for the ancestral SARS-CoV-2 spike, as well as Omicron BA.1 and BA.4/BA.5 variants. Serums were analyzed after SARS-CoV-2 infection in children (n = 26, yellow) and adults (n = 26, blue). The percentage of participants with detectable neutralizing antibodies against specific variants are indicated for each group. Statistical significance was established as : ns (not significant, not shown) P >.05, ***P<.001, ****P<.0001.

### The strength of T-cell response to SARS-CoV-2 is reduced in children compared to adults and is analogous to responses to other seasonal β−coronaviruses

Cell-mediated immune responses are critical mediators of reduced disease severity and long-term protection.^26,30,31^ Memory T-cell responses were assessed through ELISpot for the detection of IFN-𝛾 secreting cells in response to different stimulations. For all SARS-CoV-2 strains tested, children displayed less IFN-𝛾 secreting cells compared to adults seven months after SARS-CoV- 2 infection (**Fig. 3A**). The number of antigen-responding T cells per million PBMCs was two-fold lower in children compared to adults, with a median of 88 [28-184] spot forming units (SFU) per million of PBMCs in children compared to 208 [141-340] in adults (***, P < .001). There was strong immunodominance of the T-cell response towards the spike protein in children. The frequency of children displaying IFN-𝛾 secreting cells against non-Spike components (nucleocapsid and membrane protein) of SARS-CoV-2 was much lower than in adults, ****P <.0001 for both (**Fig. 3B**). The reduced magnitude of the T-cell response in children compared to adults was also observed when children who had an asymptomatic infection were excluded from the analysis (**Fig. S3**). Unexpectedly, children displayed response to SARS-CoV-2 of similar strength as they did to other seasonal β-coronaviruses, including HCoV-OC43 and HCoV-HKU1 (median of 88 [28-184] for SARS-CoV-2 compared to 58 [29-163] and 60 [33-168] SFU per million PBMCs for HCoV-OC43 and HCoV-HKU1 respectively, P >.05, not significant). In contrast, adults exhibited robust IFN-γ secretion to SARS-CoV-2 (median of 208 [141-340]) and significantly lower responses to common β-coronaviruses (median of 80 [45-135], ****P <.0001 for HCoV-OC43 compared to SARS-CoV-2 and 98 [59-151], **P <.01 for HCoV-HKU1 compared to SARS-CoV-2) (**Fig. 3C**).

**Figure 3.**
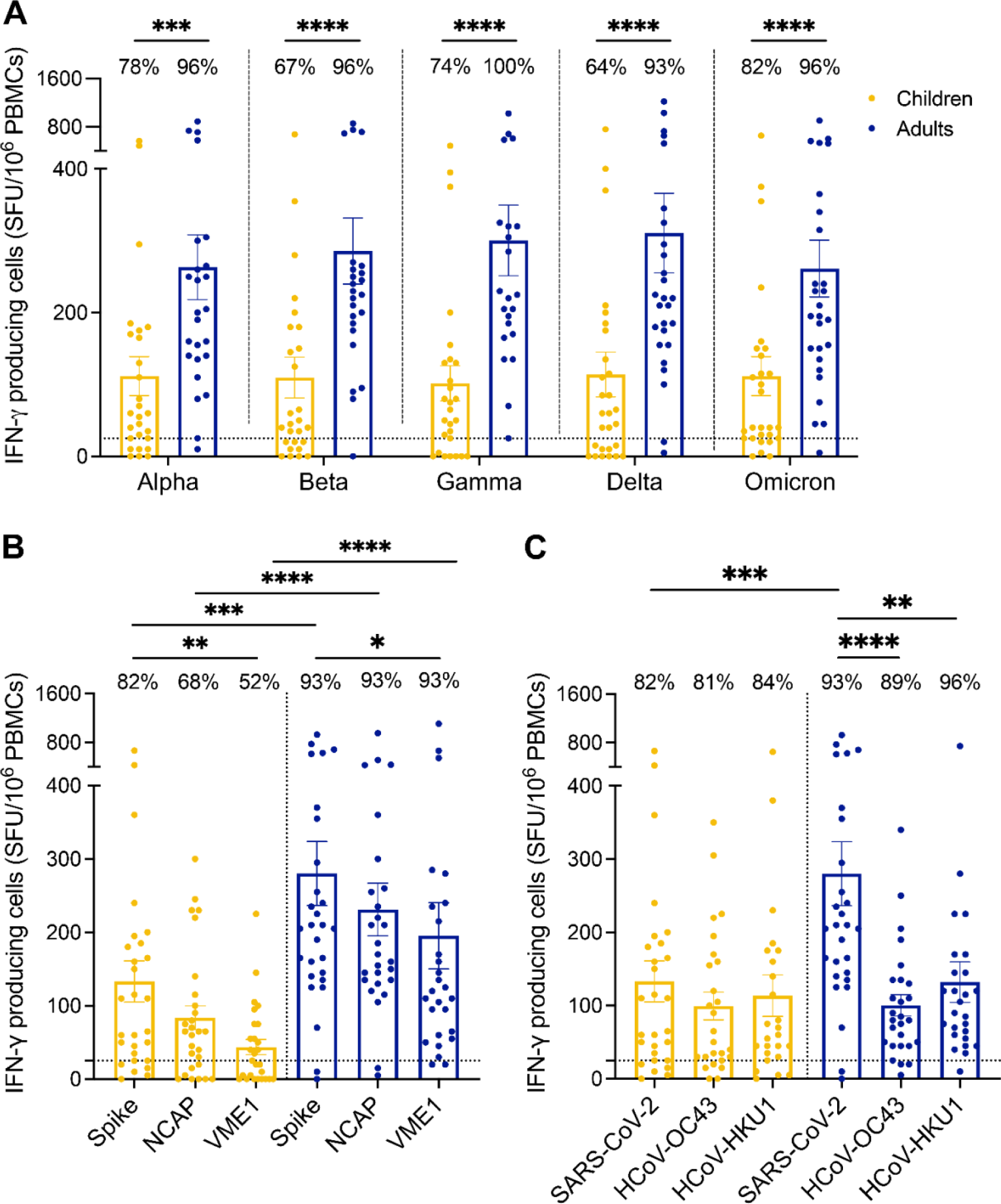
Cellular immune responses to SARS-CoV-2 are enhanced compared to other ß- coronaviruses in adults, but not in children. T-cell responses were assessed by ELISpot assay after peptide stimulation. Samples were collected from SARS-CoV-2 infected children (n = 28, yellow) and adults (n = 28, blue). **(A)** PBMCs were stimulated with SARS-CoV-2 spike peptide from five different variant strains (Alpha, Beta, Gamma, Delta, Omicron BA.1). **(B)** PBMCs were stimulated with peptide pools from the ancestral SARS-CoV-2 spike, nucleocapsid (NCAP) and membrane (VME1) protein. **(C)** PBMCs were stimulated with spike peptides from SARS-CoV-2 and common cold ß-coronaviruses HCoV- OC43 and HCoV-HKU1. Results are expressed in number of IFN-γ producing cells per million PBMCs. Dotted line indicates the positive threshold value of 25 IFN-γ secreting cells. The number of participants with responses above the positive cut-off value are indicated for each group and condition. Error bars indicate mean ± SEM. Statistical significance was established as : ns (not significant, not shown) P >.05, *P <.05, **P <.01, ***P<.001, ****P<.0001.

For all three assays used to evaluate the memory humoral and cellular responses to SARS-CoV-2 infection, no sex-based difference was observed (**Fig. S4**).

## DISCUSSION

Our study aimed to further contribute to better understanding the dynamics of the immune responses following SARS-CoV-2 infection in children. We describe the production and persistence of adaptive humoral and cellular immune responses following mild SARS-CoV-2 infection in both children and adults, with differences noted between the two groups. In the convalescence phase following acute SARS-CoV-2 infection, adults retained enhanced T-cell immune responses compared to children. In contrast, pediatric participant’s T-cell responses were lower and indistinguishable from other seasonal ß-coronaviruses.

While serum immunoglobulin levels and neutralizing capacity were comparable in both children and adults, we observed more pronounced immunodominance towards the spike versus non-spike proteins in children compared to adults at seven months post-infection. This observation is supported by other studies, who consistently report that children mount an early and effective anti-nucleocapsid response that wanes and declines more rapidly compared to adults.^10,32–34^

More importantly, we described notable differences in the intensity of the T-cell response following SARS-CoV-2 infection with lower responses in children compared to adults, when measured over six months post-infection. This observation is similar to increasing number of reports.^32,35–37^ T cells have an important role in limiting disease severity, leading to recovery but also to immune memory and protection against reinfection by future variants.^38^ Reports have shown that the severity of initial SARS-CoV-2 infection impacts the amplitude of the T-cell response, with stronger but often impaired and pathogenic responses in individuals with severe disease.^14,15,19^ Our study shows that mild disease led to higher T-cell responses to SARS-CoV-2 in adults than in children. This is consistent with the study by Khoo *et al*. who described multimodal immune responses, including cellular responses, in a cohort of five children and seven adults in the acute and convalescent phases of infection.^37^ Their results showed that adults generated stronger T-cell immune memory responses and that children showed less activation of the T-cell compartment.^37^ Our data also concurs with Cohen *et al.* who also showed that CD4^+^ memory T-cell responses to structural SARS-CoV-2 proteins increased with age.^35^ This phenomenon was suggested to be correlated to prior exposure to β-coronaviruses, with adults being exposed more frequently than children to HCoV.^35,39^ However, our data contrasts with those of Dowell *et al.*, who reported that children mount more robust adaptive humoral and cell-mediated immune responses than adults.^34^ However, their pediatric participants were younger than our cohort, which may influence the strength of the adaptive immune response.^11^ Further, recent exposure to β- coronaviruses and elevated level of HCoV cross-reactivity observed in their cohort could contribute to the differences observed between our studies. Importantly, here we show that T-cell responses to SARS-CoV-2 are strikingly elevated in adults compared to responses to OC43 and HKU1, an analysis that was not performed in the study by Dowell *et al*. Collectively, this may also highlight distinct dynamics of the immune response over time in young children compared to adults.

As mentioned above, memory T cells generated following HCoV infections are able to cross-react with SARS-CoV-2 antigens, as demonstrated using pre-pandemic samples, in both adults and children.^39–41^ This cross-reactivity is established very early in life.^39^ However, despite numerous HCoV exposures, Humbert *et al.* showed the capacity of T cells to cross-react declines with age, as does the capacity of CD4^+^ T cells to respond to HCoV.^39,41^ In line with this, we report that the increased level of T-cell memory observed in adults is unique to SARS-CoV-2 and is not seen when evaluating responses to other seasonal HCoV, as demonstrated by their lower level of response to HCoV-HKU1 and HCoV-OC43. In our study, children exhibited cellular responses to SARS-CoV-2 comparable to that elicited in response to other seasonal ß-coronaviruses. This is interesting as the clinical picture of SARS-CoV-2 for the majority of children is also very similar to other HCoV. It may suggest that SARS-CoV-2 leads to inherently distinct immune responses in adults and children, with SARS-CoV-2 being recognized as would other seasonal coronaviruses in children.

The differences between adults and children in the strength of the T-cell responses to SARS-CoV- 2 could be explained by a rapid and extremely efficient innate response in children. Indeed, it is thought that children have pre-activated and robust innate immune responses that are very effective at controlling and clearing SARS-CoV-2 infection.^17,42^ This would greatly impact the secondary T-cell response as early viral clearance by the innate immune response would restrict antigen availability and the amplitude of long-lived T-cell responses.^17,43^ Further, an accelerated innate immune response may lead to a more coordinated and balanced adaptive immune response in children, which could contribute to their milder symptoms and reduced inflammatory complications.

Although there is general homogeneity in the clinical presentation of the cases, there are limitations to our study, mostly related to sampling. The time from infection to blood sampling was variable between study participants, ranging from two to fifteen months post-infection (median of seven months). Blood sampling was also limited due to pediatric volume restrictions. Lastly, due to the limited cell numbers, peptide or protein-specific mapping was not possible. The main strength of our study is the direct comparison of both humoral and cell-mediated immunity following mild SARS-CoV-2 infection in adults and children, also interrogating responses to seasonal coronaviruses. Collectively, our results suggest that in the long-term convalescent phase, T-cell responses to SARS-CoV-2 are attenuated in children compared to adults and are similar to that of other common seasonal coronaviruses.

## CONCLUSION

Previous studies suggest that robust innate immune responses help to clear SARS-CoV-2 infection rapidly in children, similar to other seasonal coronaviruses. This highly effective clearance of viral antigens may lead to less T-cell activation, therefore inducing a distinct T-cell differentiation program and dampened IFN-γ^+^ T-cell responses in children compared to adults. Further longitudinal studies are needed to establish the diversity and duration of protection mediated by T cells in children and whether vaccination is useful in broadening and increasing the durability of protection against SARS-CoV-2 infection and severe disease in children.

## Data Availability

All data produced in the present study are available upon reasonable request to the authors.

## ACKNOWLEDGMENTS

We first want to recognize and warmly thank all of the participants and their parents/guardians who agreed to participate in these studies, as well as the nurses and research coordinators from each participating centers. We are also very grateful for the Children’s Hospital of Eastern Ontario, the Coronavirus Variants Rapid Response Network (CoVaRR-Net) Biobank and the Sainte-Justine University Hospital Mother Child Biobank personnel for processing and storing the samples from the studies. Special thanks to Candice McGahern, Fazia Tadount, Kelsey Adams, Louise Wang and Sylvie Nicholson for their role in participant recruitment and sampling as well as Angela Crawley, Laura Tamblyn, Jocelyne Ayotte, Jessie Beauchemin, Vanessa Truong, Annie Bilodeau and Amal Abdi for their role in sample preparation. ELISA assays were performed by the University of Ottawa Serology and Diagnostics High Throughput Facility. Special thanks to Danielle Dewar-Darch, Justino Hernandez-Soto, Abishek Xavier, Kiran Nakka, Smita Upamaka and Yuchu Dou. COVID-19 antigens (variant S trimer and RBD proteins), as well as the anti-hIgG#5-HRP fusion antibody were generously provided by Dr. Yves Durocher, National Research Council of Canada (NRC), Montréal.

## Author contributions

Conceptualization: RLZ, MB, HD, APH

Methodology: SN, CA, YG, BB, RLZ, MAL, MB, HD, APH

Investigation: HD, APH Visualization: SN, BB, HD

Funding acquisition: RLZ, CQ, MB, HD, APH Project administration: BB, JB

Subject recruitment: JB, CQ, APH Supervision: HD, APH

Writing – original draft: SN, BB, HD, APH

Writing – review & editing: SN, CA, BB, JB, RLZ, CQ, MB, HD, APH

## ABBREVIATIONS

ACE2: Angiotensin-converting enzyme 2
COVID-19: Coronavirus disease 2019
DMSO: Dimethyl sulfoxide
ELISA: Enzyme-linked immunosorbent assay
ELISpot: Enzyme-linked immunospot
HCoV: Human Coronavirus
IFN-γ: Interferon-gamma
IgA: Immunoglobulin
A IgG: Immunoglobulin G
IQR: Interquartile range
nAb: Neutralizing antibody
N: Nucleocapsid
PBMCs: Peripheral blood mononuclear cells
RBD: Receptor-binding domain
RPMI media: Roswell Park Memorial Institute media
SARS-CoV-2: Severe acute respiratory syndrome coronavirus 2
SFU: Spot forming units
snELISA: Surrogate neutralisation enzyme-linked immunosorbent assay
SOPs: Standard operating procedures
S: Spike

