## Supplementary Material for "Differential Adaptive Immune Responses Following SARS-CoV-2 Infection in Children Compared to Adults"

**Running Title : SARS-CoV-2 Immune Responses : Are Children Less Protected ?**

**Authors :** Sabryna Nantel,[1,2] Corey Arnold,[3] Maala Bhatt,[4,5] Yannick Galipeau,[3] Benoîte Bourdin,[1] Jennifer Bowes,[4] Roger L. Zemek,[4,5] Marc-André Langlois,[3] Caroline Quach,[1,2] Hélène Decaluwe [1,2,6,#,*] & Anne Pham-Huy [4,7,#,*]

**Affiliations :**

[1] Sainte-Justine University Hospital and Research Center, Montréal, Québec, Canada.

[2] Department of Microbiology, Infectious Diseases and Immunology, Faculty of Medicine, University of Montréal, Montréal, Québec, Canada.

[3] Department of Biochemistry, Microbiology and Immunology, Faculty of Medicine, University of Ottawa, Ottawa, Ontario, Canada.

[4] Children's Hospital of Eastern Ontario Research Institute, University of Ottawa, Ottawa, Ontario, Canada.

[5] Division of Emergency Medicine, Department of Pediatrics, University of Ottawa, Ottawa, Ontario, Canada.

[6] Pediatric Immunology and Rheumatology Division, Department of Pediatrics, University of Montréal, Montréal, Québec, Canada.

[7] Division of Infectious Diseases, Immunology and Allergy, Department of Pediatrics, Children's Hospital of Eastern Ontario, University of Ottawa, Ottawa, Ontario, Canada.

### Shared senior authorships

**CORRESPONDING AUTHORS (*)**

Hélène Decaluwe

CHU Sainte-Justine Research Center

3175, Chemin de la Côte-Sainte-Catherine, Montréal, QC, Canada (H3T 1C5)

Anne Pham-Huy

Children's Hospital of Eastern Ontario

401, Smyth Road, Ottawa, ON, Canada (K1H 8L1)

#### SUPPLEMENTAL FIGURES AND LEGENDS

##### Supplemental Figures

**Figure S1.** The IgA response against the nucleocapsid remains lower when comparing only symptomatic children to adults.

**Figure S2.** Symptomatic children present slightly increased neutralizing antibody titers to Omicron BA.4/BA.5 compared to symptomatic adults.

**Figure S3.** The reduced cellular immune response to SARS-CoV-2 in children compared to adults is still observed after excluding asymptomatic children.

**Figure S4.** The immune response to SARS-CoV-2 is similar in both sex.

**Figure S1. The IgA response against the nucleocapsid remains lower when comparing only symptomatic children to adults.**

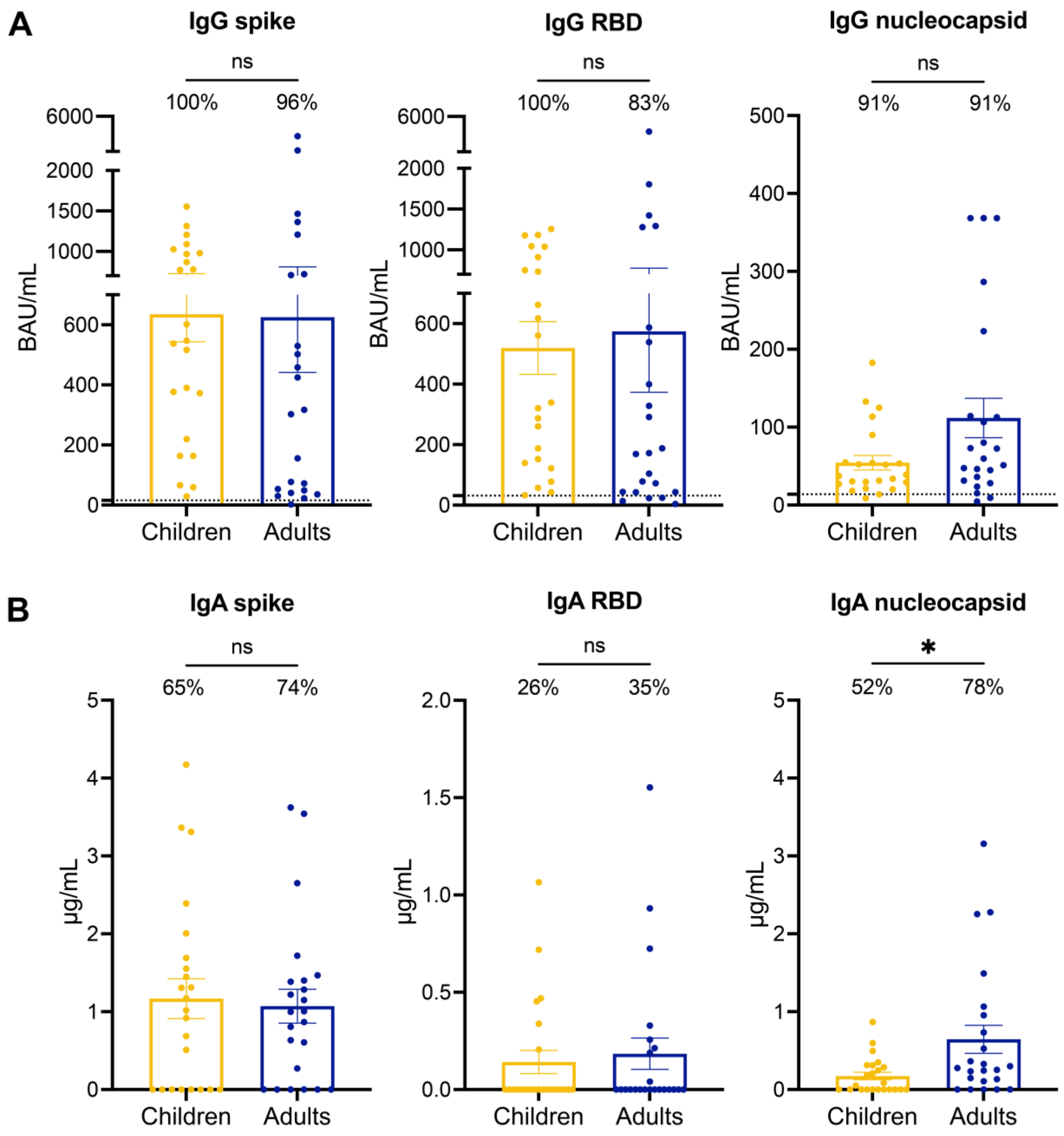


ELISA were conducted to measure the anti-spike **(left panels)**, anti-RBD **(middle panels)** and anti-nucleocapsid **(right panels)** IgG **(A)** and IgA **(B)** levels in the serum of symptomatic and seropositive children (n = 23, yellow) and adults (n = 23, blue) infected with SARS-CoV-2. **(A)** The percentage of participants with IgG responses above the positive cut-off value (15.53216 for spike, 31.34807 for RBD and 13.83556 for nucleocapsid) are indicated for each group. **(B)** The percentage of participants with detectable IgA are indicated for each group. Error bars indicate mean ± SEM. Statistical significance was established as : ns (not significant) $P > .05$, $*P < .05$.

**Figure S2. Symptomatic children present slightly increased neutralizing antibody titers to Omicron BA.4/BA.5 compared to symptomatic adults.**

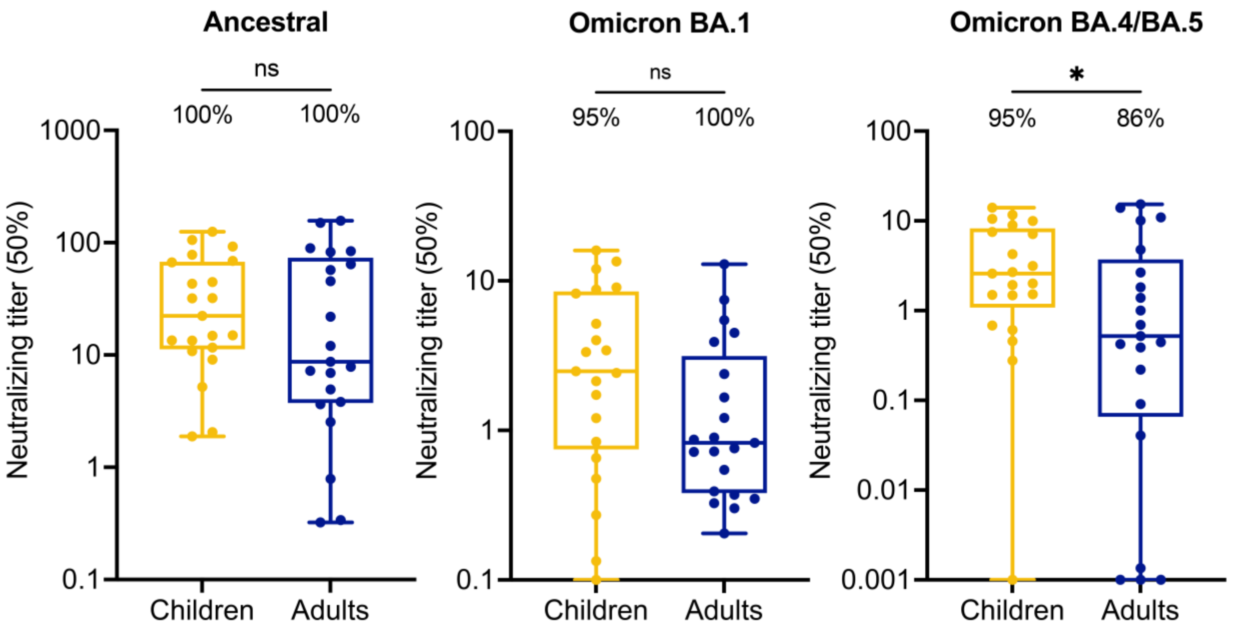


Surrogate neutralization ELISA were conducted to establish the serum dilution level (ID50) required to inhibit 50% of the binding between the trimeric spike protein and the ACE2 receptor, thus neutralizing the virus's attachment capability to cells. Neutralizing antibody titers were measured for the ancestral SARS-CoV-2 spike, as well as Omicron BA.1 and BA.4/BA.5 variants. Serums were analyzed after SARS-CoV-2 infection in symptomatic and seropositive children (n = 21, yellow) and adults (n = 21, blue). The percentage of participants with detectable neutralizing antibodies against specific variants are indicated for each group. Statistical significance was established as : ns (not significant) P >.05, *P <.05.

**Figure S3. The reduced cellular immune response to SARS-CoV-2 in children compared to adults is still observed after excluding asymptomatic children.**

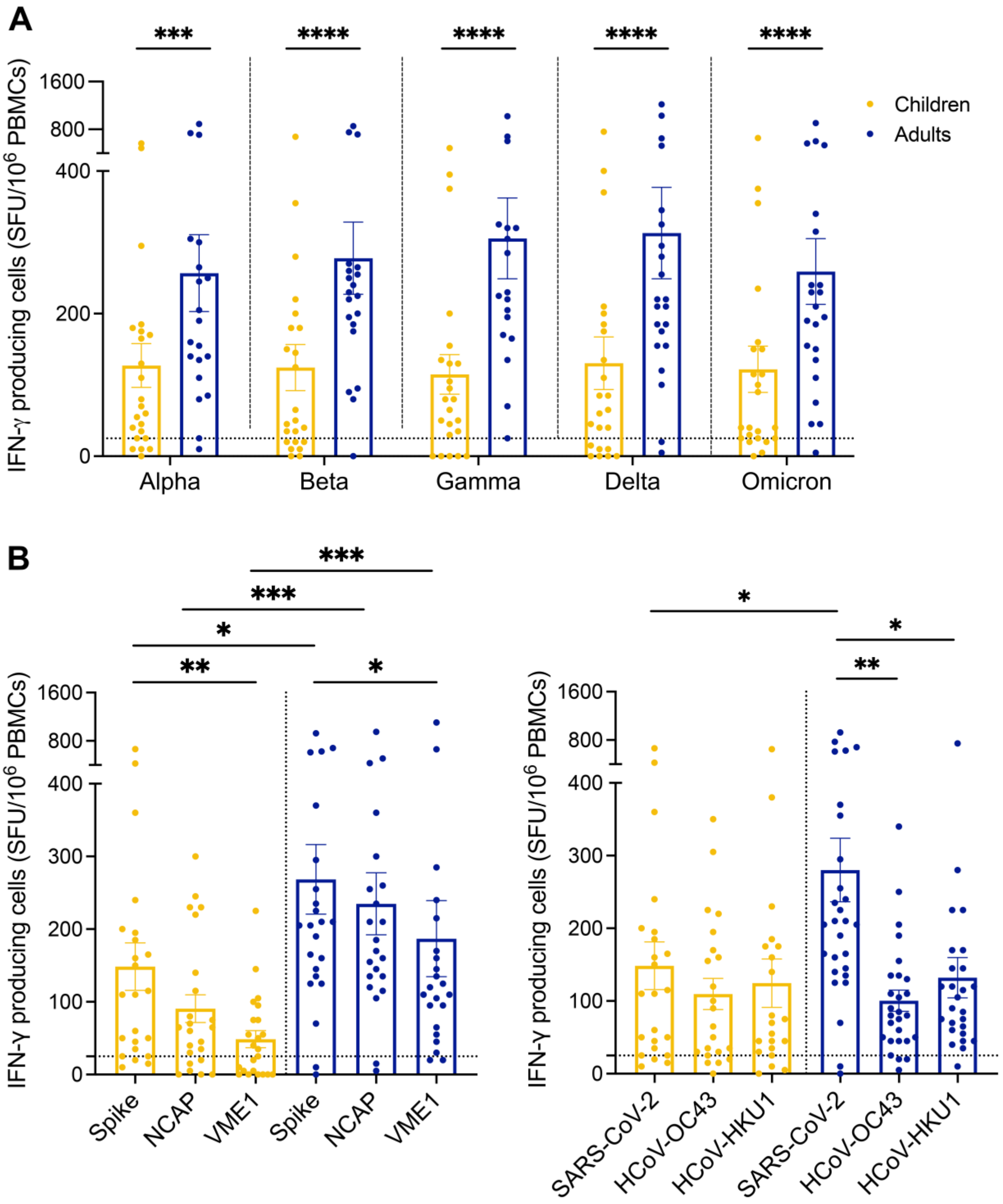

T-cell responses were assessed by ELISpot assay after peptide stimulation. Samples were collected from SARS-CoV-2 infected children who were symptomatic at the time of infection, and seropositive at sampling (n = 23, yellow) and adults (n = 23, blue). **(A)** PBMCs were stimulated with SARS-CoV-2 spike peptide from five different variant strains (Alpha, Beta, Gamma, Delta, Omicron BA.1). **(B)** PBMCs were stimulated with peptide pools from the ancestral SARS-CoV-2 spike, nucleocapsid (NCAP) and membrane (VME1) protein. **(C)** PBMCs were stimulated with spike peptides from SARS-CoV-2 and common cold ß-coronaviruses HCoV-OC43 and HCoV-HKU1. Results are expressed in number of IFN-γ producing cells per million PBMCs. Dotted line indicates the positive threshold value of 25 IFN-γ secreting cells. Error bars indicate mean ± SEM. Statistical significance was established as : ns (not significant, not shown) P >.05, *P <.05, **P <.01, ***P<.001, ****P<.0001.

**Figure S4. The immune response to SARS-CoV-2 is similar in both sex.**

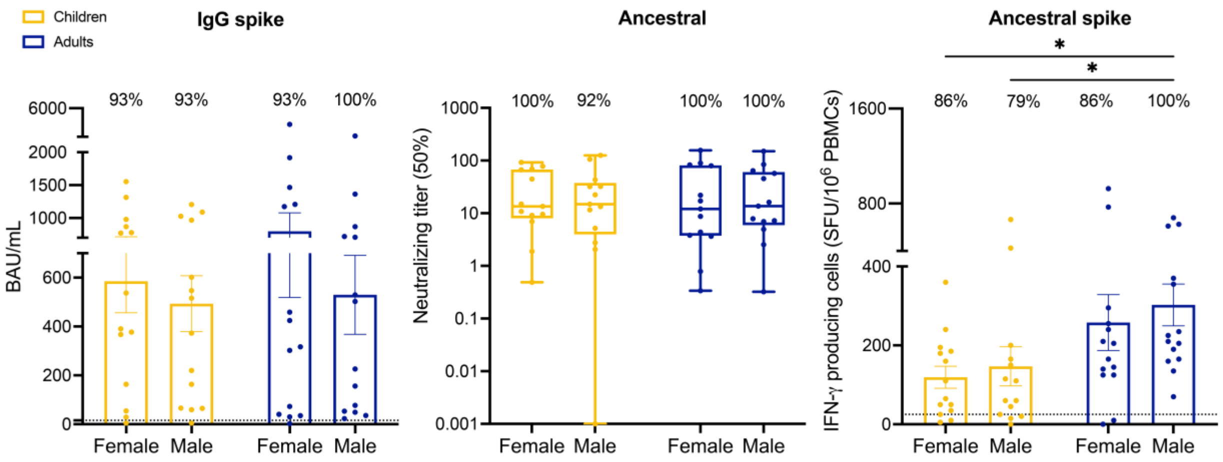


Samples were collected from equal numbers of children (n = 28) and adults (n = 28) from each sex (n = 14). **(A)** ELISA were conducted to measure the anti-spike IgG in the serum. **(B)** Surrogate neutralization ELISA were conducted to establish the serum dilution level (ID50) required to inhibit 50% of the binding between the trimeric spike protein and the ACE2 receptor, thus neutralizing the virus attachment capability to cells. **(C)** T-cell responses were assessed by ELISpot assay after peptide stimulation. For each of the three assays performed, no sex-based difference were noted. Error bars indicate mean ± SEM. Statistical significance was established as : ns (not significant) P >.05, *P <.05.
